# Quality controlled SARS-CoV-2 duplex procedure to reduce time and scarce molecular biology diagnosis reagents

**DOI:** 10.1101/2020.07.28.20154153

**Authors:** Annaelle Collin, Carole Chaboteaux, Véronique Fontaine, Philippe Lefèvre

## Abstract

The aim of this study was to provide validated procedures allowing to detect the SARS-CoV-2 and an internal control in a unique one-step duplex RT-qPCR. Two internal controls were tested, targeting either the Schmallenberg virus RNA provided by the NARILIS laboratory (University of Namur) with a HEX-labelled probe or a Diagenode Diagnostics internal control with a Cy5-labelled probe. Our results showed that Ct values of the RT-qPCR duplex assay were even smaller in the optimized working conditions, allowing to use the optimized qPCR conditions in routine diagnosis.

## INTRODUCTION

A current international health crisis is shaking the world due to the new severe acute respiratory syndrome coronavirus 2 (SARS-CoV-2) pandemic infection causing the coronavirus disease 2019 (COVID-19). The burden is not only on the health care system but also on our society putting our economy at risk. Although most infected individuals (approximately 80 %) show benign clinical signs, vulnerable and older people can show severe symptoms [1]. Amongst the various option to reduce the ongoing outbreak, rapid and early diagnosis of symptomatic and asymptomatic infected individuals are highly appreciated to rapidly quarantine the SARS-CoV-2 infected one’s or to rapidly clinically enroll patient with severe symptoms. As the SARS-CoV-2 is a positive single-stranded RNA virus, virology laboratories can routinely detect with high sensitivity and specificity its viral genome present in various clinical samples by reverse-transcriptase qualitative real-time polymerase chain reaction (RT-qPCR). A rapid researcher solidarity allowed to share not only genomic information (Wuhan-Hu-1, GenBank accession number MN908947) deposed on the Global Initiative on Sharing All Influenza Data (GISAID) but also in-house developed laboratory assay protocols to detect and identify the emerged coronavirus deposed among others on the WHO website [2, 3].

Based on their previous experiences, the team of C. Drosten together with an European research network, published in January 2020 a validated diagnosis workflow to detect the SARS-CoV1 [4]. Using Taqman probes in reverse-transcriptase - quantitative polymerase chain reaction (RT-qPCR), detection of the envelop protein (E) sequence allowed for a highly sensitive pan-coronavirus detection tool, with an analytical sensitivity of up to 3.2 viral RNA copies per reaction, more specifically the new SARS-CoV-2 genome [4].

In the absence of physical sources of positive control SARS-CoV-2 genomic sequence, quality controls of the clinical laboratory assays rely on documentation of their analytical qualification and validation.

The impressive ongoing deployment of laboratory assay capacity by coordinated action of academic, public and private laboratories is however highly dependent of reagent availability. Indeed rapid lacks of specific commercial kits seriously impeded clinical laboratory diagnosis capacity. Rapidly, the additional support of fundamental research laboratories allowed to develop quality controlled diagnosis workflows. In Belgium, a veterinary research unit of the University of Namur (NARILIS) developed and provides an internal control, RNA extract containing Schmallenberg virus RNA genome, for each RT-qPCR reaction [5]. SARS-CoV-2 RT-qPCR are consequently only validated when a parallel RT-qPCR targeting the Schmallenberg virus RNA genome show positive results. Similar internal controls are also commercially available. Those controls allow to assess nucleic acid extraction quality and to verify the absence of PCR inhibitor or process error. Although their sequences have no similarities with the SARS-CoV-2 sequences, they are usually performed in time and reagent consuming parallel Taqman RT-qPCR assays, further increasing the risk of detection kit and reagent shortage [5].

The aim of this study was to provide validated procedures allowing to detect the SARS-CoV-2 and an internal control in a unique one-step duplex RT-qPCR.

## MATERIALS AND METHODS

### Clinical specimen collection

Nasopharyngeal swabs (N=15) were obtained during the March 2020 and May 2020 from patients requiring SARS-CoV-2 molecular biology screening test from the Clinical Microbiology laboratory of the Hospital Princesse Paola of Marche-en-Famenne (IFAC– VIVALIA).

### RNA extraction

RNA was extracted from the clinical samples by the QIAamp Virus Pathogen Midi Kit on a QIASymphony automate (QIAGEN, Hilden, Germany) using manufacturer’s intructions. Before RNA extraction procedure, an internal control was added to the clinical samples, consisting either in 5 μL Schmallenberg virus RNA extract or in 10 μL Cy5 (DRIC-CY-L100) RNA control.

### Real-time reverse transcription PCR

Reactions to detect SARS-CoV-2 E sequence were performed in 25 μL as described by Coupeau *et al*. [5]. Briefly, a 25 μL reaction contained RNAse free water, 5 μL 5X concentrated reaction master mix containing DNA polymerase, MgCl_2_ and dNTP (Eurogentec Takyon One-Step No Rox Probe 5X MasterMix dTTP), 0.25 μL Euroscript II reverse transcriptase and RNAse inhibitor and 0.25 μL additive (both provided with the Eurogentec Takyon 5X MasterMix), 5 μL RNA and various concentration of primers and probes. In singleplex assays, primers and probes (targeting either the SARS-CoV-2 E or the Schmallenberg virus RNA) were at 400 nM and 200 nM final concentration, respectively, as previously described [5]. In duplex assays, the working concentration of primers and probe targeting the SARS-CoV-2 E sequence were at 100 nM and 50 nM, respectively, and the working concentration of primers and probe targeting the Schmallenberg virus RNA internal control were at 200 nM and 100 nM, respectively. Alternatively, when we used the Cy5 internal control of Diagenode Diagnostics (Seraing, Belgium), 0.5 μL of RNA internal control targeting Cy5 primers and probe (DICR-CY-L100) were included in the 25 μL reaction. Thermocycler reactions were performed in the Rotor-Gene Q (QIAGEN, Hilden, Germany) according to the following program : 10 min. 48°C, 95°C 3 min. followed by 45 cycles of 15 sec. at 95°C and 30 sec. at 58°C, as previously described. Fluorescence were measured at 510 nm and 555 nm when using Schmallenberg virus RNA internal control and additionally at 660 nm when using Diagenode Diagnosis Cy5 internal control.

### Statistical analysis

The independent t student test for paired samples was used to verify the significance between the various Ct values obtained for same RNA samples in three RT-qPCR methods, and using the singleplex RT-qPCR as reference. This was performed using the Graphpad tools available on https://www.graphpad.com/quickcalcs/ttest2/.

## RESULTS AND DISCUSSION

In order to reduce time and reagent consumption, we worked to achieve the best qPCR reaction conditions allowing to detect SARS-CoV-2 positive patient samples in a duplex one-step RT-qPCR (testing various master mixes and various primer and probe concentrations). This was performed using residues of various positive sample RNA extracts remaining after the routine singlexplex one-step RT-qPCR. The most satisfying conditions were further validated using fifteen new RNA samples among the various RNA specimens collected for suspicion of SARS-CoV-2 infection using nasopharyngeal swabs. They were selected to be part of one of the 3 subgroups containing 5 RNA samples (Ct25, Ct30 or Ct35) based on a singleplex one-step RT-qPCR Ct result being around 25, 30 or 35, respectively. The mean of the Ct values obtained by the singleplex one-step RT-qPCR for these positive specimens were 25.01, 30.36 and 36.47 for the FAM SARS-CoV-2 probe (Figure 1). Using the conditions described under “Materials and Methods”, we obtained satisfying Ct value means for the FAM SARS-CoV-2 probe in the duplex one-step RT-qPCR using the Schmallenberg virus RNA extract as internal control (24.43, 29.57, 37.33, respectively) or using the Cy5 Diagenode Diagnosis internal control (24.99, 30.04, 34.16, respectively) (Figure 1). Statistical analysis of the Ct value means showed that the Ct values obtained in the duplex one-step RT-qPCR with the Cy5 Diagenode Diagnosis internal control were particularly satisfying to detect the SARS-CoV-2 RNA in samples with low viral load. Indeed, for Ct around 35, the p value in the independent t student test for paired samples was significatively different with a p value of 0.0315, a 95% confidence interval (CI) from 0.3353 to 4.2967 and a standard error of difference (SED) of 0.713. In case of higher viral load (Ct values around 25 or 30), the duplex one-step RT-qPCR with the Cy5 Diagenode Diagnosis internal control performed similarly to the simplex RT-qPCR (for samples with Ct around 25, the p= 0.8299, 95% CI is -0.2000 to 0.2360 and the SED is 0.138; for samples with Ct around 30, the p= 0.0716, 95% CI is -0.0452 to 0.6892 and the SED is 0.132) suggesting that the duplex one-step RT-qPCR with the Cy5 Diagenode Diagnosis internal control could be used in order to reduce time and reagent consumption. When performing the duplex one-step RT-qPCR with the Schmallenberg virus RNA extract as internal control, although this duplex one-step RT-qPCR could statistically perform better when using high viral load (for Ct around 25, the p = 0.0006, 95% CI is 0.4114 to 0.7406 and SED is 0.059; for Ct around 30, the p = 0.0046, 95% CI is 0.4058 to 1.1702 and the SED is 0.138), the Ct value difference were quite reduced. Considering that in the presence of low viral load the duplex one-step RT-qPCR with the Schmallenberg virus RNA extract as internal control could also perform as the singleplex (p = 0.5024, 95%CI = -4.0931 to 2.3771, SED = 1.165), the one-step RT-qPCR could be perform in duplex to save time and reagents.

**Figure 1.**
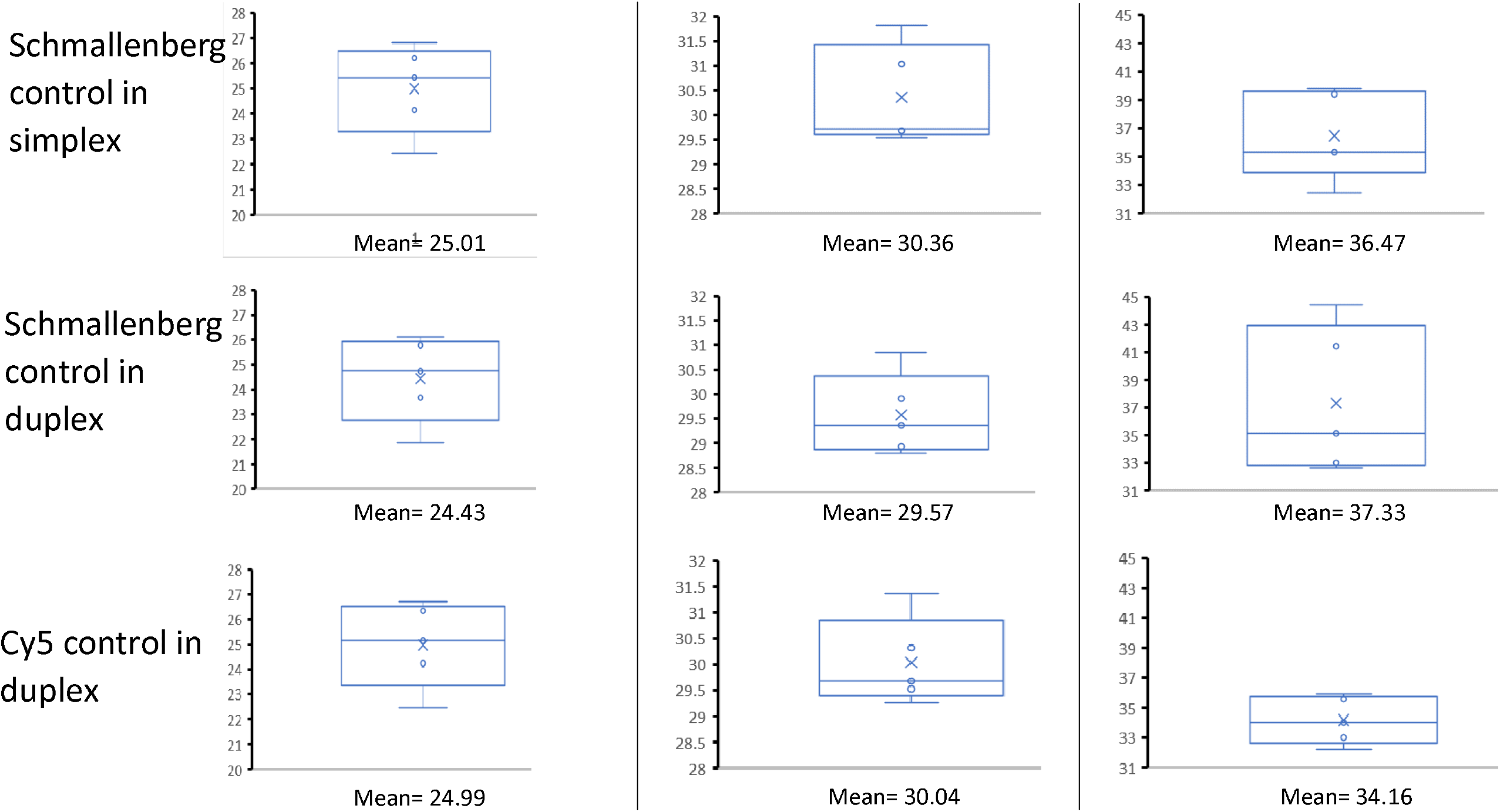
Ct values of the FAM SARS-CoV-2 probe in the singleplex (simplex) one-step RT-qPCR reaction (used as reference for the statistical analysis), or in the duplex one-step RT-qPCR reaction using the Schmallenberg virus RNA extract as internal control or in the duplex one-step RT-qPCR reaction using the Cy5 Diagenode Diagnosis as internal control.

## CONCLUSION

To save time and reagents, we recommend to perform a duplex one-step RT-qPCR reaction using either the Schmallenberg virus RNA extract or the Cy5 Diagenode Diagnosis as internal controls, using the conditions described in the Materials and Methods.

## Data Availability

the datasets used and analyzed during the current study are de-identified and available from the corresponding author on reasonable request

